# Assessing different next-generation sequencing technologies for wastewater-based epidemiology

**DOI:** 10.1101/2024.05.22.24306666

**Authors:** Anika John, David Dreifuss, Seju Kang, Anna Bratus-Neuenschwander, Natalia Zajac, Ivan Topolsky, Arthur Dondi, Catharine Aquino, Timothy R. Julian, Niko Beerenwinkel

## Abstract

Wastewater-based epidemiology has proven to be an important public health asset during the COVID-19 pandemic. It can provide less biassed and more cost-effective population-level monitoring of the disease burden as compared to clinical testing. An essential component of SARS-CoV-2 wastewater monitoring is next-generation sequencing, providing genomic data to identify and quantify circulating viral strains rapidly. However, the specific choice of sequencing method influences the quality and timeliness of generated data and hence its usefulness for wastewater-based pathogen surveillance. Here, we systematically benchmarked Illumina Novaseq 6000, Element Aviti, ONT R9.4.1 MinION flow cell, and ONT R9.4.1 Flongle flow cell sequencing data to facilitate the selection of sequencing technology. Using a time series of wastewater samples from influent of six wastewater treatment plants throughout Switzerland, along with spike-in experiments, we show that higher sequencing error rates of ONT Nanopore sequencing reduce the accuracy of estimates of the relative abundance of viral variants, but the overall trend is in good concordance among all technologies. We find that the sequencing runtime for ONT Nanopore flow cells can be reduced to as little as five hours without significant impact on the quality of variant estimates. Our findings suggest that SARS-CoV-2 variant tracking is readily achievable with all tested technologies, albeit with different tradeoffs in terms of cost, timeliness and accuracy.

## 1. Introduction

During the recent COVID-19 pandemic, wastewater-based epidemiology (WBE) has emerged as a significant public health resource as it offers a less biassed and more cost-efficient method for monitoring disease prevalence at the population level compared to clinical testing^1–4^. Next-generation sequencing (NGS) plays a crucial role in SARS-CoV-2 wastewater monitoring by enabling tracking of the prevailing SARS-CoV-2 variants in communities. The effectiveness of wastewater monitoring has further been shown in several studies by demonstrating its ability to detect emerging variants of concern earlier than clinical surveillance^5–7^. Different NGS technologies have found application in wastewater monitoring as they provide unique advantages for WBE.

Illumina sequencing provides high coverage and is considered one of the best NGS technologies in terms of sequencing accuracy^8^. Hence it is frequently applied in comparative studies between wastewater and clinical surveillance^5–7^. Illumina sequencing is a sequencing by synthesis method, where fluorescently labelled nucleotides are incorporated sequentially followed by an imaging step.

A recent competitor of the well-established Illumina sequencing is the Aviti sequencing platform. Through the use of a novel sequencing chemistry, incorporation and identification of nucleotides are optimised, resulting in higher sequencing accuracy than Illumina^9^. Nevertheless, Aviti sequencing has not yet been applied in the context of WBE to the best of our knowledge.

Nanopore-based sequencing, which is mainly distributed by Oxford Nanopore Technologies (ONT), provides a cost-efficient alternative to Illumina and Aviti sequencing and is the sequencing method of choice in several wastewater studies, including the ARTIC network wastewater surveillance efforts^10–13^. In contrast to Illumina sequencing, Nanopore sequencing is not synthesis-based but instead nucleotides pass through a polymer membrane via a nanoscale protein pore (Nanopore) and are typed through the disruption of an electric current across the membrane. The entire sequencing reaction is integrated on a single flow cell, which can contain thousands of Nanopores. Different ONT flow cells exist, which vary in throughput and sequencing accuracy^14^. In this study we focused on the ONT MinION R9.4.1 and Flongle R9.4.1 flow cells and will refer to them as ‘Nanopore flow cells’ throughout the paper. The small size of these flow cells and the development of equally compact sequencing instruments, provide portable sequencing options^15^. This portability and the underlying molecular process allows for real-time monitoring and dynamic adjustment of sequencing runtime^16^. As Nanopore flow cells can be reused by subjecting them to a washing protocol, dynamic runtime adjustment can further reduce sequencing costs^17^. Thus, Nanopore sequencing is also attractive for settings where reduced cost and lower burden on laboratory infrastructure are crucial factors for a successful implementation of wastewater surveillance. The advantages of Nanopore sequencing come at the cost of lower sequencing quality^16,18^ as compared to Illumina sequencing.

One of the main demonstrated applications of NGS for wastewater monitoring in the context of the COVID-19 pandemic is to estimate the relative abundances of different SARS-CoV-2 variants in samples over time^6,7,19–21^. To this aim, computational tools are used for deconvolving the observed mutation frequencies in the sample into variant relative abundances, i.e., for finding the relative abundances of variants that best explain the observed pattern of mutation frequencies^6,19–21^. Tracking the relative abundance of variants through time and geographical locations can provide prime epidemiological information. In this study we evaluate the effect of sequencing wastewater samples with different technologies on the ability to accurately deconvolve the mutation data into the relative abundances of variants.

Since NGS methods influence quality and timeliness of pathogen surveillance in wastewater, here we provide a systematic assessment of Illumina Novaseq 6000, Element Aviti, ONT MinION flow cell, and ONT Flongle flow cell sequencing to facilitate the selection of sequencing method based on the study’s objectives. Our comprehensive evaluation incorporates time-resolved wastewater samples from six wastewater treatment plants (WWTPs) across Switzerland along with spike-in experiments.

## 2. Methods

### 2.1 Sampling and processing of wastewater

Surveillance of SARS-CoV-2 through wastewater analysis was carried out in six sewersheds (Altenrhein, Chur, Laupen, Lugano, Geneva, Zurich) across Switzerland from January 4, 2023, to January 10, 2023. Daily composite samples, obtained from 24-hour flows of wastewater influent at each of the six locations, were collected and preserved at 4°C for a maximum of 5 days. Subsequently, these samples were transported on ice to a central laboratory (Eawag, Dübendorf, Switzerland) for processing. The processing steps involved total nucleic acid extraction from 40 ml samples by using the Wizard Enviro Total Nucleic Acid Extraction Kit (CN A2991, Promega Corporation, USA) with an elution volume of 80 µl as described in Nadeau et al.^22^. Following this, inhibitor removal was performed by using OneStep PCR Inhibitor Removal columns (CN D6030, Zymo Research, USA). All samples (n = 36) were subsequently subjected to the sequencing protocols described below. RNA extracts were stored at −80°C for up to one week prior to sequencing. The wastewater sequencing data of all NGS technologies is available on the European Nucleotide Archive (ENA) under project accession number PRJEB44932^23^.

### 2.2 Spike-in experiment

For the BA.1 Omicron strain spike-in experiment, the clinical Omicron strain (BA.1) was obtained from clinical isolates confirmed as BA.1 via sequencing, obtained from the Functional Genomic Center Zurich. Its concentration was measured to be 25.1 gc/µL by quantifying the N1 gene target by using Crystal Digital PCR (Naica system, Stilla Technologies) as described in the previous study ^7^. In parallel, as a wild-type solution, twenty RNA extracts from SARS-CoV-2 Omicron-free wastewater (August 2021, the same six sewersheds across Switzerland) were pooled together. The pooled solution was quantified to be 36.5 gc/µL of wild-type SARS-CoV-2 by using the same digital PCR assay targeting the N1 gene.The mixtures of wild-type and BA.1 solutions were prepared for the BA.1 percentage to be 88.3, 77.5, 57.9, 49.1, 40.7, 18.6, 8.9, 4.4, 2.2, 1.1 and 0 %, with respective concentrations 22.17, 17.17, 9.94, 4.88, 1.99, 0.37, 0.03, 1.45 × 10^-3^, 3.4 × 10^-7^ and 0 gc/µL. Each mixture was sequenced in triplicates with the sequencing platforms described below. The sequencing data can be found here https://doi.org/10.3929/ethz-b-00066382724.

### 2.3 Library preparation for Illumina sequencing

The RNA extracts were reverse transcribed using the LunaScript Super Mix (New England Biolabs, Franklin Lake, NJ, USA) according to manufacturer’s instructions. The resulting cDNA were amplified using the ARTIC V4.1 NCOV-2019 Panel (IDT, Iowa, USA). Sequencing libraries were prepared using the NEBNext Ultra II Prep Kit for Illumina (New England Biolabs, Franklin Lake, NJ, USA) and were used in the succeeding steps according to manufacturer’s instructions. Briefly, 2.5 µl of PCR products (cDNA amplicons) were end-repaired before ligation of adapters. Fragments containing adapters on both ends were selectively enriched with PCR implementing unique dual indices (UDI) for multiplexing. The quality and quantity of the enriched libraries were validated using a TapeStation (Agilent, Santa Clara, California, USA). The product is a peak of a size of approximately 500 bp. For each sample, 5 of the libraries were used for pooling.

### 2.4 Illumina sequencing

The pooled libraries were further quality checked using a TapeStation (Agilent, Santa Clara, California, USA) and quantified with Qubit HS DNA assay (Thermofisher Scientific, USA) and were loaded into a NovaSeq 6000 (Illumina, Inc, California, USA), 18µl of the pooled libraries with a concentration of 0.8 nM was loaded on a lane of a NovaSeq 6000 SP Reagent Kit v1.5 (500 cycles) flow cell for a final loading concentration of 180 pM. The wastewater sequencing data of all NGS technologies is available on the European Nucleotide Archive (ENA) under project accession number PRJEB44932^23^.

### 2.5 Aviti sequencing

The pool of Illumina libraries was prepared for sequencing on the AVITI sequencer (Element Biosciences, San Diego, CA) using the Element Adept Library Compatibility Kit v1.1. This process involves the denaturation, library circularization via ligation to a splint adapter, and exonuclease digestion of non-circularized molecules. Thirty microliters of the Illumina sequencing library pool at a concentration of 16.7 nM were circularised. The resulting circularised library was quantified via qPCR using the standards provided in the compatibility kit. Twenty-five microliters of the circularised library at a concentration of 3.5 pM sequenced with a AVITI 2×300 Sequencing Kit. The wastewater sequencing data of all NGS technologies is available on the European Nucleotide Archive (ENA) under project accession number PRJEB44932^23^.

### 2.6 ONT sequencing

Viral cDNA amplicons (400 bp long) were produced in a tiled fashion across the whole SARS-CoV-2 genome following the SARS-CoV-2 ARTIC V4.1 NEB Ultra II protocol (the same as for Illumina sequencing). The Oxford Nanopore libraries were further produced from cDNA amplicons by ligation of native barcodes (Native Barcoding Expansion 96 (EXP-NBD196)) and subsequent sequencing adapter ligation (Ligation Sequencing Kit (SQK-SQK109) following the instruction from Oxford Nanopore Technologies protocols: ‘pcr-tiling-SARS-CoV-2-nbd-PTCN_9103_v109_revQ_13Jul2020-minion’ and ‘amplicon-barcoding-with-native-barcoding-expansion-96-exp-nbd196-and-sqk-NBA_9102_v109_revN_09Jul2020-minion’.

Each Oxford Nanopore library was sequenced on both MinION flow cell (R9.4.1 (FLO-MIN106) and Flongle flow cell (R9.4.1 (FLO-FLG001)) on the ONT Mk1C device with 72h and 24h run time, respectively. The wastewater sequencing data of all NGS technologies is available on the European Nucleotide Archive (ENA) under project accession number PRJEB44932^23^.

### 2.7 Effect of the sequencing technology on lineage relative abundance estimation in the spike-in data

To determine whether technology or runtime have an effect on the estimates of relative abundances, we modelled the relationship between the relative abundances of the variant Omicron BA.1 determined from the dilution *f* and its relative abundance estimated from deconvolution of the sequencing data *f*’. We modelled this relationship as a linear function after applying the arcsine square root transformation, which is commonly used for proportional data as it is the variance-stabilising transformation for binomial likelihoods ^25^,

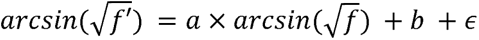

Where *a* and *b* are the slope and intercept parameters, respectively, and *∈* is a random error term. To account for the presence of heteroskedasticity in the residual errors and hence increase the robustness of our model, we additionally calculate the error variance using the “Huber Sandwich Estimator” ^26,27^. We included fixed additive effects of the experimental conditions (technology or runtime) on the parameters *a* and *b*. The model was fitted by using robust linear regression implemented in *stats* (R base package). The statistical significance of the effects of technology or runtime on the parameters *a* and *b* were assessed by using two-sided t-tests. When comparing the different sequencing technologies, the model was fitted multiple times with different treatment contrasts to allow for all pairwise comparisons of technologies (no correction for multiple comparisons was applied). When comparing different runtimes for the sequencing, the model was fitted with treatment contrasts set with the lowest sequencing time (5h) as a reference level. The code is available here https://doi.org/10.5281/zenodo.1108572128.

### 2.8 Bioinformatic pre-processing

Basecalling and demultiplexing was performed by using Guppy (version 6.4.6) integrated in the MinKnow software (version 22.12.5). The “Fast model, 450 bps’’ basecalling model was applied. To adhere to community standards pre-processing was subsequently performed by using the wf-artic workflow ^29^, with parameters `--normalise 200, --_min_len 400, --_max_len 700 and --_scheme_version ARTIC/V4.1`. Version 1.3.0 of the arctic pipe-line was used. Other parameters were set to default values. Alignment was performed with minimap2 (version 2.18-r1015), with -x map-ont set. Using the information provided in the sequencing summary file, reads were sub-sampled from the bam files depending on the sequencing time point by using the samtools view -bS -N command.

### 2.9 Variant abundance estimation

The alignment files resulting from the pre-processing described above were analyzed by using V-pipe (version 3.0)^30^, a bioinformatics pipeline for analysis of viral sequencing reads obtained from mixed samples, which has been adapted for wastewater sequencing data. By using the integrated tool LolliPop^31^ SARS-CoV-2 signature mutations were deconvolved in lineage relative abundances for the variants of interest (B.1.1.7, B.1.351, P.1, B.1.617.2, B.1.617.1, BA.1, BA.2, BA.4, BA.5, BA.2.75, BQ.1.1 and XBB). The kernel bandwidth parameter was set to 1e-17. Detailed documentation of the analysis settings can be found here https://doi.org/10.5281/zenodo.1108572128.

### 2.10 Sequencing error rate estimation

Sequencing error rates were determined from clinical Omicron (BA.1) isolates, with an expected BA.1 relative variant abundance of 1.0. BA.1 signature mutations were defined as single nucleotide polymorphisms present uniquely in BA.1 and not in earlier reported variants. Hence we included mutations listed in Table 1 (relative to reference NC_045512.2). The sequencing error rate was defined as the mean fraction of reads lacking the signature mutation. This resulted in position-wise error rates of which the mean was taken.

**Table 1:** BA.1 unique signature mutations relative to NC_045512.2 reference sequence.

| Genomic position | Mutation |
| --- | --- |
| 2832 | A to G |
| 5386 | T to G |
| 8393 | G to A |
| 11537 | A to G |
| 13195 | T to C |
| 15240 | C to T |
| 21762 | C to T |
| 21846 | C to T |
| 23048 | G to A |
| 23202 | C to A |
| 24130 | C to A |
| 24503 | C to T |
| 26530 | A to G |

## 3. Results

We investigated the suitability of the different sequencing technologies for wastewater based SARS-CoV-2 variant abundance estimation by using two separate experimental setups (Figure 1). First, we compared variant abundance estimates from a real life setting by using 42 wastewater samples originating from six geographical locations in Switzerland, sampled daily for one week in January 2023^32^. In Swiss clinical samples from the same time period, BQ.1, BA.5 and BA.275 were the most abundant circulating variants. The surveillance samples, collected within the Swiss wastewater surveillance program, underwent identical treatment up to library preparation under the ARTIC protocol, and were subsequently sequenced by using Illumina Novaseq 6000, Element Aviti, one R9.4.1 MinION flow cell, and one R9.4.1 Flongle flow cell. The second experiment compared variant abundance estimates obtained from the above-mentioned sequencing methods in samples with a predefined combination of SARS-CoV-2 variants. Through spike-in experiments we simulated the arrival of a novel SARS-CoV-2 variant (BA.1) and analysed deviations from expected abundances for the different sequencing technologies. For nanopore sequencing data we also investigated the effect of sequencing runtime on the variant abundance estimates. For both experiments, the sample processing and relative abundance estimation of SARS-CoV-2 variants was performed using V-pipe^30^, a workflow designed for the analysis of NGS data from viral pathogens. The experimental setup and bioinformatics data analysis is described in detail in Methods.

**Figure 1:**
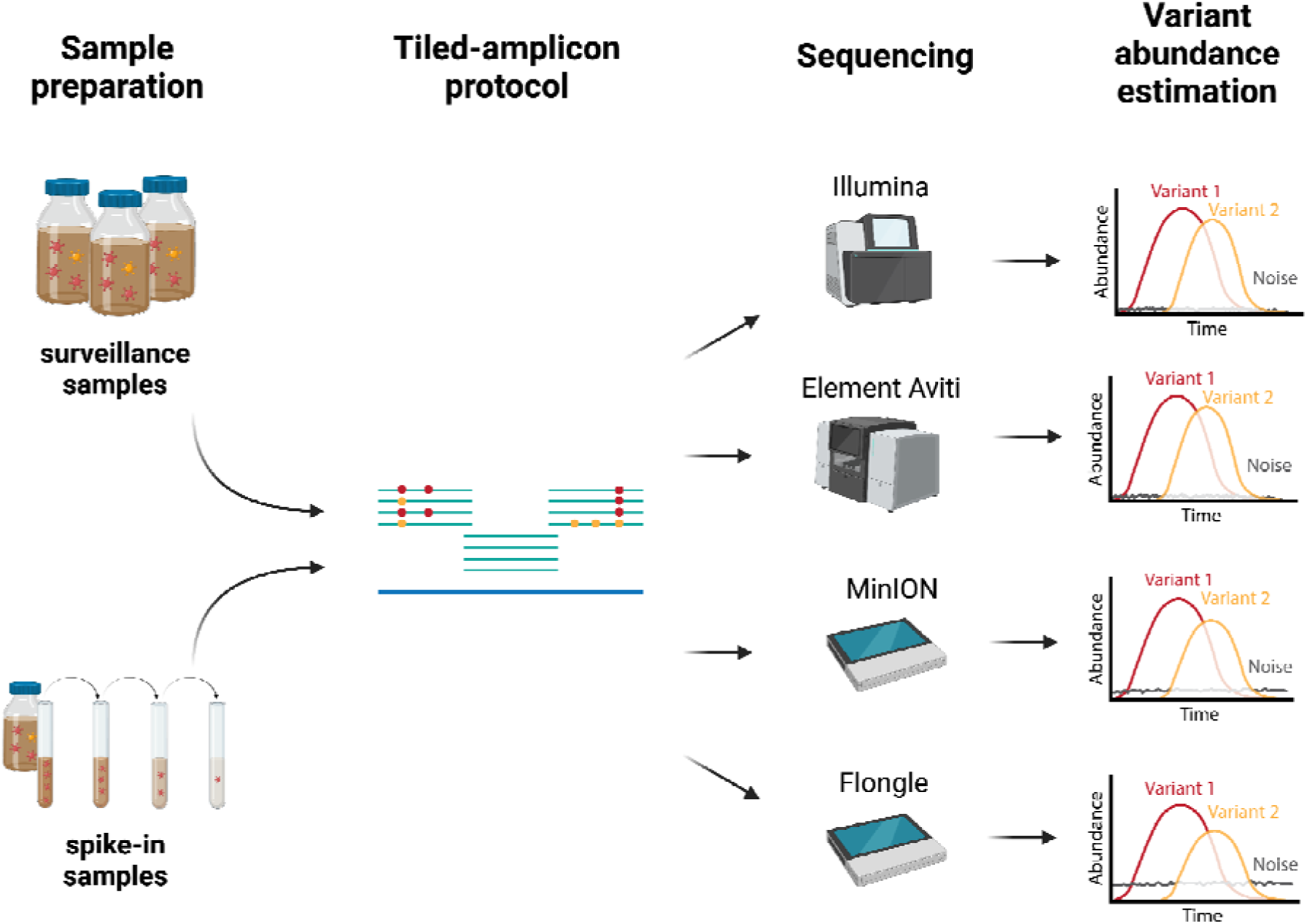
Overview. One week of daily collected wastewater samples from six WWTPs was subjected to RNA extraction and SARS-CoV-2 v.4.1 ARTIC tiling-amplicon amplification. The resulting cDNA samples were sequenced with Illumina Novaseq 6000, Element Aviti, and Oxford Nanopore flow cells (MinION and Flongle) technologies. Next the sequencing data was used as input for the bioinformatics analysis to estimate relative abundance of different viral variants. These results were then statistically compared across sequencing technologies. The Figure was created with BioRender.com.

### 3.1 Whole-genome coverage is achieved for all sequencing technologies

For the surveillance samples we obtained the highest mean amplicon coverage using Illumina sequencing (Figure 2A), with coverage drops caused by PCR amplification biases during amplicon generation. The most prominent drops are visible around amplicons 14 and 73, where mean amplicon coverages approach zero. Despite being lower, the viral genome coverages obtained with the Aviti, MinION and Flongle flow cells follow a similar overall pattern to the Illumina coverage (Figure 2A). Notably, the apparently more uniform coverage of MinION flow cell data originates from a coverage normalisation step during pre-processing of the data (section 2.8). Aviti sequencing data also shows very similar coverage compared to Illumina sequencing data (Figure 2B). The consistent amplicon coverage pattern for all sequencing technologies indicates an absence of technology-specific sequencing bias across amplicons.

**Figure 2:**
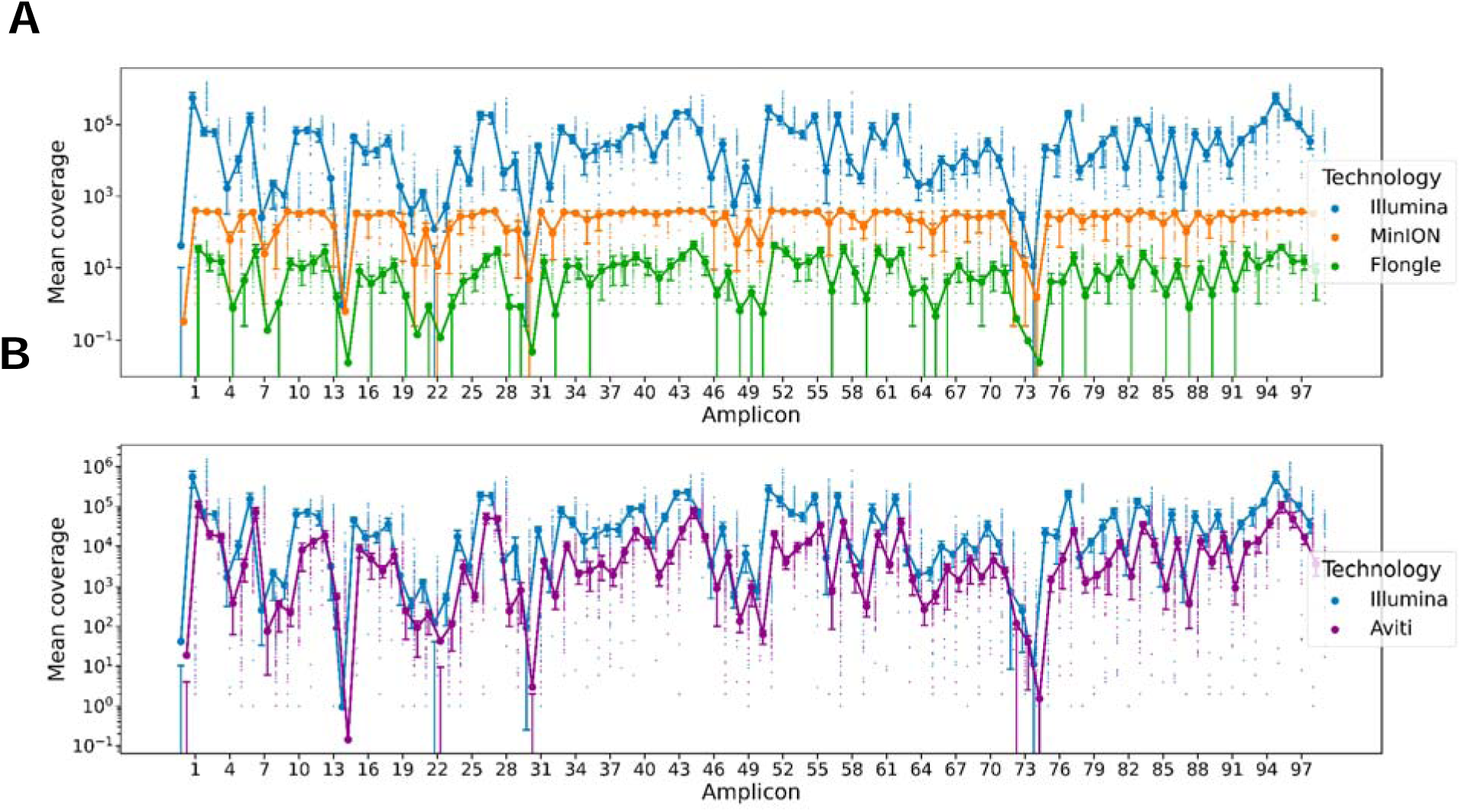
Mean amplicon coverage distribution for the sequencing technologies of interest amongst 42 surveillance samples. The amplicons for all sequencing technologies were generated by using v4.1 ARTIC primers. For each amplicon the mean coverage over all 42 samples is represented by a large dot. The smaller dots represent the individual coverage depth of each amplicon within one of the 42 samples. The error bar at each amplicon represents the interquartile range of the coverage values (pi = 50). An overview of the amplicon distribution and coverage depth across the genome is shown in Supplementary Figure A. **A:** Comparing amplicon coverage between Illumina- and Nanopore-based surveillance samples. **B:** Comparing amplicon coverage between Illumina- and Aviti-based surveillance samples.

### 3.2 Nanopore sequencing displays higher error rates compared to Illumina and Aviti

To estimate the sequencing error rate for each sequencing technology, we assessed the respective deviation of 13 BA.1 signature mutation frequencies from BA.1 clinical isolates with expected relative abundance of 1.0 (see section 2.10). We found a mean sequencing error rate of 0.0038 for Illumina, 0.0032 for Aviti, 0.1 for MinION and 0.2 for Flongle sequencing. The Nanopore technologies also showed a larger spread in error rate compared to Illumina and Aviti, with Flongle data displaying the largest spread (Figure 3). The position-wise error rate shows a similar distribution across positional coverage (Supplementary Figure B).

**Figure 3:**
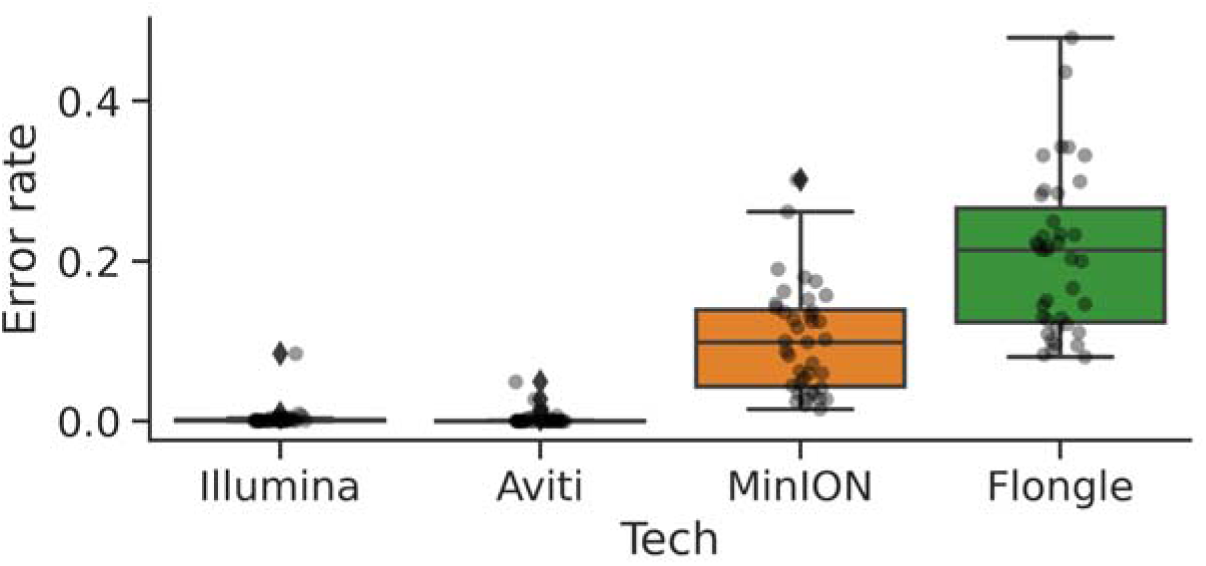
Boxplots of per-position error rates. For each sequencing dataset, error rates were estimated from the three non-diluted spike-in samples, where the expected relative abundance of BA.1 is 1.0. Only the positions of the 13 BA.1 signature mutations were considered. For more details see section 2.10. The box of the boxplots represents the interquartile range and the line within the box represents the median of the values. The whiskers of the box plots extend to points that lie within 1.5 times the upper and lower quartile. Values outside this range are displayed as rhombus. The transparent dots represent the individual per-position error rates.

### 3.3 Variant abundance estimates from Nanopore and Aviti technologies correlate well with Illumina-based estimates

For the 42 surveillance samples we determined the mutation frequencies of SARS-CoV-2 variant signature mutations (Figure 4A). Subsequently we estimated the relative abundance of SARS-CoV-2 variants of interest, for each sample separately by deconvolution of signature mutation data using V-pipe (Figure 4B). For each sequencing technology we compared the results to the outcomes obtained from Illumina data, as it was the first technology in the context of the monitoring program.

**Figure 4:**
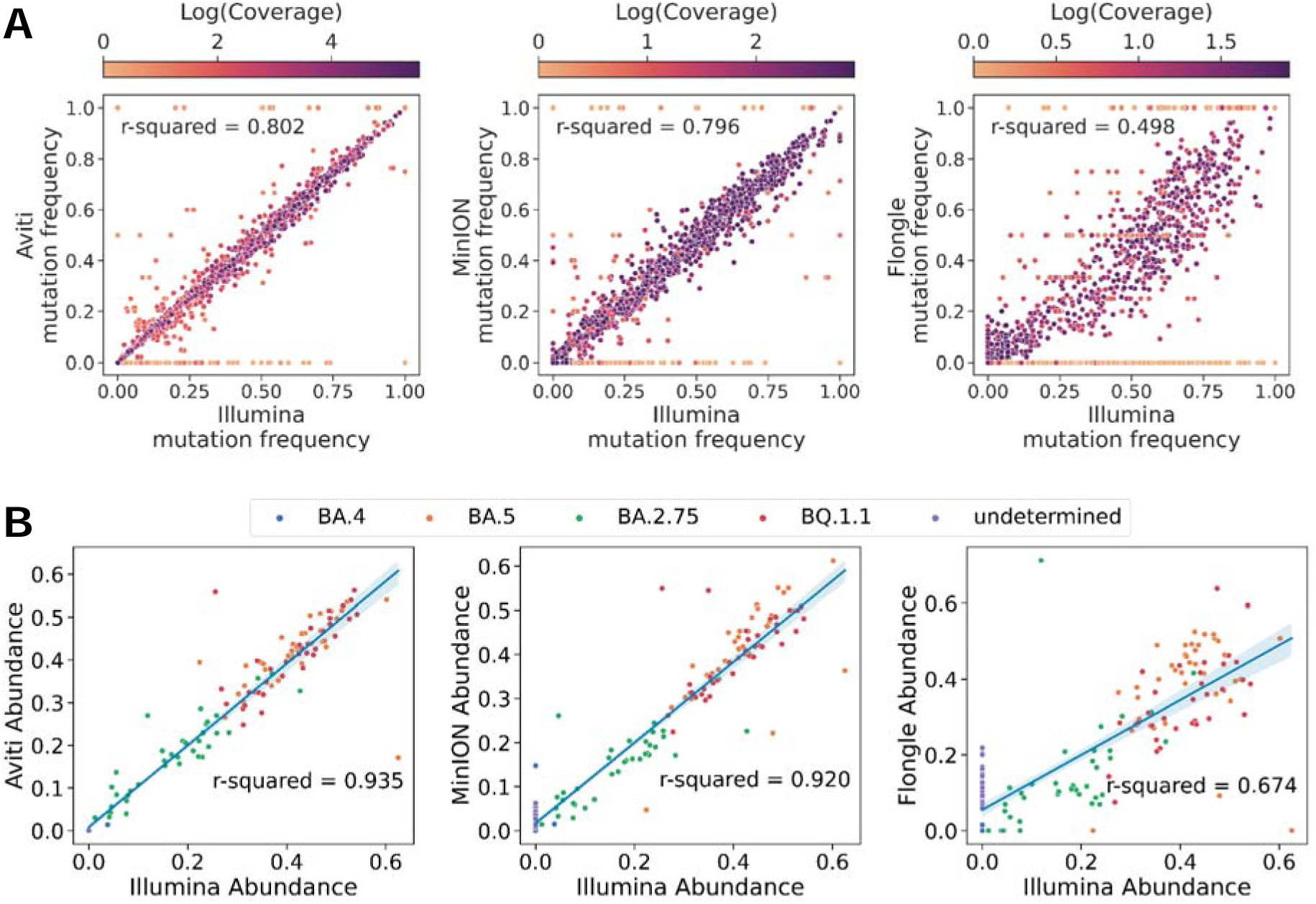
Comparison of mutation frequencies and variant abundance estimations by sequencing technologies compared to Illumina. **A:** Each dot represents a unique mutation for a SARS-CoV-2 variant of interest for one of the 42 surveillance samples included in this plot. Dots are coloured by the log(coverage) of y-axis-based sequencing technology at the respective mutation position. Dark purple corresponds to a high coverage, light yellow to a low coverage. **B:** Each dot represents the estimated abundance of a given variant for one of the 42 surveillance samples included in this plot.The abundances were estimated by using LolliPop. Lines represent the estimated linear regression on the points and the shaded area the corresponding 95% confidence intervals. Mutations that cannot be classified to a defined variant, are categorised as “undetermined” by LolliPop.

Considering SARS-Cov-2 signature mutations (Figure 4A), we observed the largest deviation from Illumina-based mutation frequencies for the Flongle data (R^2^ = 0.498, p-value < 2.2e^-16^). For the Aviti and MinION data, deviations from the Illumina-based mutation frequencies were observed, but less prominent (Aviti: R^2^ = 0.802, p-value < 2.2e^-16^; MinION: R^2^ = 0.796, p-value < 2.2e^-16^) and mostly associated with low coverage (Supplementary Figure C). The BA.2.75 unique signature mutations frequently displayed low coverage, since a substantial amount of them is located on amplicon 73 and 74, which experienced a coverage drop (Figure 2). For the Flongle data large deviations were also observed for moderately covered positions (Supplementary Figure D).

The results observed when comparing variant abundance estimates across technologies (Figure 4B) aligned with the previous results from the mutation frequency correlation analysis. We found that overall, variant abundance estimates from Nanopore sequencing correlated well (MinION: R^2^ = 0.920, p-value < 2.2e^-16^; Flongle: R^2^ = 0.674, p-value < 2.2e^-16^) with Illumina-based estimates (Figure 4B). However, estimates obtained from Flongle data displayed a larger spread (Figure 4B), especially for the BA.5 variant, (Supplementary Figure E). Further, the higher abundance of the undetermined variant category (assigned in the presence of non-signature mutations) in the Flongle data suggests the frequent occurrence of undefined mutations within the Flongle data set. These findings further supported the indication of a higher sequencing error rate for Flongle sequencing.

Variant abundance estimates obtained from Aviti data were most similar to Illumina-based estimates (R^2^ = 0.935, p-value < 2.2e^-16^) across the majority of variants of interest (Figure 4B and Supplementary Figure B). The mutation frequency observations and the absence of undetermined variants (Figure 4B) provide additional evidence for a lower sequencing error rate of Aviti compared to the Nanopore technologies. Further we found that the most prominent outliers from the Illumina-Aviti abundance correlation (Figure 4B) corresponded to samples which have a lower overall coverage in the Illumina dataset (Supplementary Figure F). Such an outlier pattern is not evident for the Nanopore data (Supplementary Figure F) and hence indicated that deviations in Illumina- and Aviti estimates are most likely due to differences in coverage.

### 3.4 MinION and Flongle flow cell data show no significant difference in spike-in BA.1 variant abundance estimates

We assessed the difference between sequencing technologies further through a spike-in experiment where a known concentration of the BA.1 variant was diluted in a wastewater background sample containing mainly B.1.617.2 (see section 2.2). Twelve dilution steps were performed in three replicates, and samples were sequenced by using Illumina Novaseq 6000, Element Aviti, one MinION flow cell, and one Flongle flow cell. The resulting sequencing reads were processed and SARS-CoV-2 signature mutations deconvolved in lineage relative abundances as for the surveillance samples under 3.3 (see section 2.9). We then systematically compared the expected concentration of BA.1 to the estimates obtained from the different sequencing technologies (Figure 5). To assess the effect of the sequencing technology on the variant abundance estimation, we modelled the estimated relative abundances as a linear response to the relative abundances expected from the mixing concentrations and defined sequencing technology as a qualitative factor of the model (see section 2.7).

**Figure 5:**
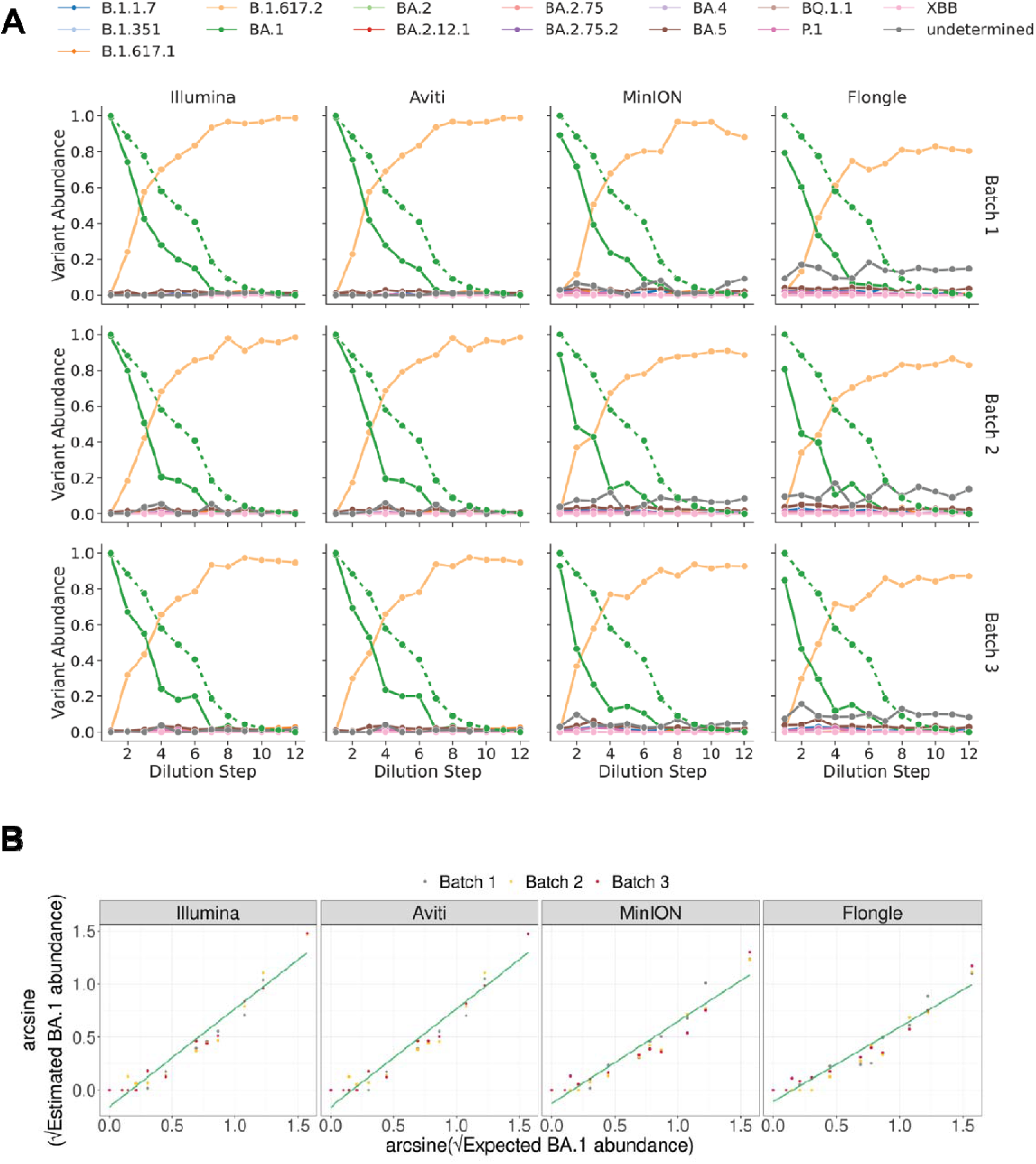
Variant Abundance estimation from spike-in experiment. Abundance estimations were made by using LolliPop. For each sequencing technology, three technical replicates were provided. **A:** Each row represents the abundance estimates for one technical replicate (batch). The green dotted line shows the expected BA.1 abundance, for ‘Dilution Step’ 1 represents the BA.1 clinical isolate (see section 2.2). **B:** Comparison of expected and estimated BA.1 abundance on arcsine-square root scale. The green line represents the derived linear model (see section 2.7).

When comparing relative variant abundance of BA.1 across sequencing technologies, we observed a high correlation between Aviti-, Nanopore- and Illumina-based BA.1 abundance estimates. In addition good concordance of whole-genome amplicon coverage was present across all sequencing technologies (Supplementary Figure G).

Consistent with the comparison of sequencing technologies on the surveillance data (Figure 4), we observed more noise (constant low-level detection of spurious variants) in estimates from Nanopore flow cell data, with Flongle data displaying the highest level of noise (Figure 5A). Although deviations from the expected abundances were clearly visible for all sequencing technologies (Figure 5A), they followed the same trend across technologies. Averaging the frequencies of BA.1-unique signature mutations (Supplementary Figure H), produced similar results compared to Figure 5A, indicating that the deviation from the expected abundance is also present in the initial sequencing reads and not introduced through variant abundance estimation. Hence the divergence between expected and estimated BA.1 abundance was most likely caused by preparation of the spike in samples in the lab, such as inaccurate initial quantification of SARS-CoV-2 RNA using digital PCR; inaccurate pipetting; or RNA degradation during the spike-in.

To assess the effect of sequencing technology on variant abundance estimation we modelled the relative abundance of the BA.1 variant estimated from the deconvolution of the sequencing data as a linear function (on the arcsine square root scale, variance-stabilising transformation for binomial likelihoods) of the expected relative abundance of BA.1 from the dilution series (see section 2.2). We found no significant difference between the slopes (p-value = 0.98, two-sided t-test) and intercepts (p-value = 0.96, two-sided t-test) of the response curves from Illumina and Aviti data (Figure 5B). Similarly, we found no significant difference between the slopes (p-value = 0.23, two-sided t-test) and intercepts (p-value = 0.6, two-sided t-test) of the response curves from MinION and Flongle flow cell data (Figure 5B, Supplementary Table A). However, our model predicted a significant difference between the slopes of the Nanopore-derived response curves compared to Illumina-(MinION: p-value = 2.183 × 10^-2^, Flongle: p-value = 2.846 × 10^-4^; two-sided t-test) and Aviti-derived (MinION: p-value = 2.060 × 10^-2^, Flongle: p-value = 2.602 × 10^-4^; two-sided t-test) response curves (see Supplementary Table A and B). This indicated a statistically significant underestimation of BA.1 for Nanopore-derived data (the slope parameter of MinION and Flongle differs to the Illumina slope parameter by 18% and 27% respectively).

Next, we checked whether the underestimation of BA.1 for Nanopore-derived data, is caused by the higher noise levels or instead by other sources of bias that might be sequencer specific. To do so we re-normalized the relative abundance of BA.1 and B.1.617.2 variant estimates, through re-calculating the relative abundance of BA.1 by considering only BA.1 and B.1.617.2 to be present in the variant mix. Doing so we removed low-level spurious variants (noise) from the data. Fitting this re-normalized data to the same model, we no longer observed a significant difference between the slopes of the Nanopore-derived response curves compared to the Illumina (MinION: p-value = 0.32, Flongle: p-value = 0.26; two-sided t-test) and Aviti (MinION: p-value = 0.31, Flongle: p-value = 0.25; two-sided t-test) response curves (see Supplementary Table C, Table D, and Figure I), indicating that the underestimation of BA.1 was due to higher noise levels with Nanopore.

### 3.5 Reducing Nanopore sequencing runtime to five hours has no significant impact on BA.1 abundance estimation

As Nanopore sequencing by ONT allows for real-time sequencing, we investigated the impact of sequencing runtime on variant abundance estimates, for the two ONT flow cells. For MinION flow cell sequencing, a large proportion of good quality sequencing reads was generated within the first five to ten hours of the sequencing run (Figure 6A, Supplementary Tables E). For the Flongle flow cell, even the majority of good quality sequencing reads was generated during the first ten hours of the sequencing run (Supplementary Tables F). In order to investigate whether reducing the sequencing runtime to this time span has a significant effect on BA.1 variant abundance estimation, we downsampled the reads to represent 15 h, 10 h, and 5 h of sequencing time, from which we produced estimates of the relative abundance of variants as described previously. We again modelled the relationship between estimated and expected variant abundances and incorporated runtime as a factor of the model (see section 2.7). We found that reducing the runtime to as little as five hours does not have a significant effect on the slope and intercept of the response curve for either flow cell (Figure 6B, Supplementary Table E and F).

**Figure 6:**
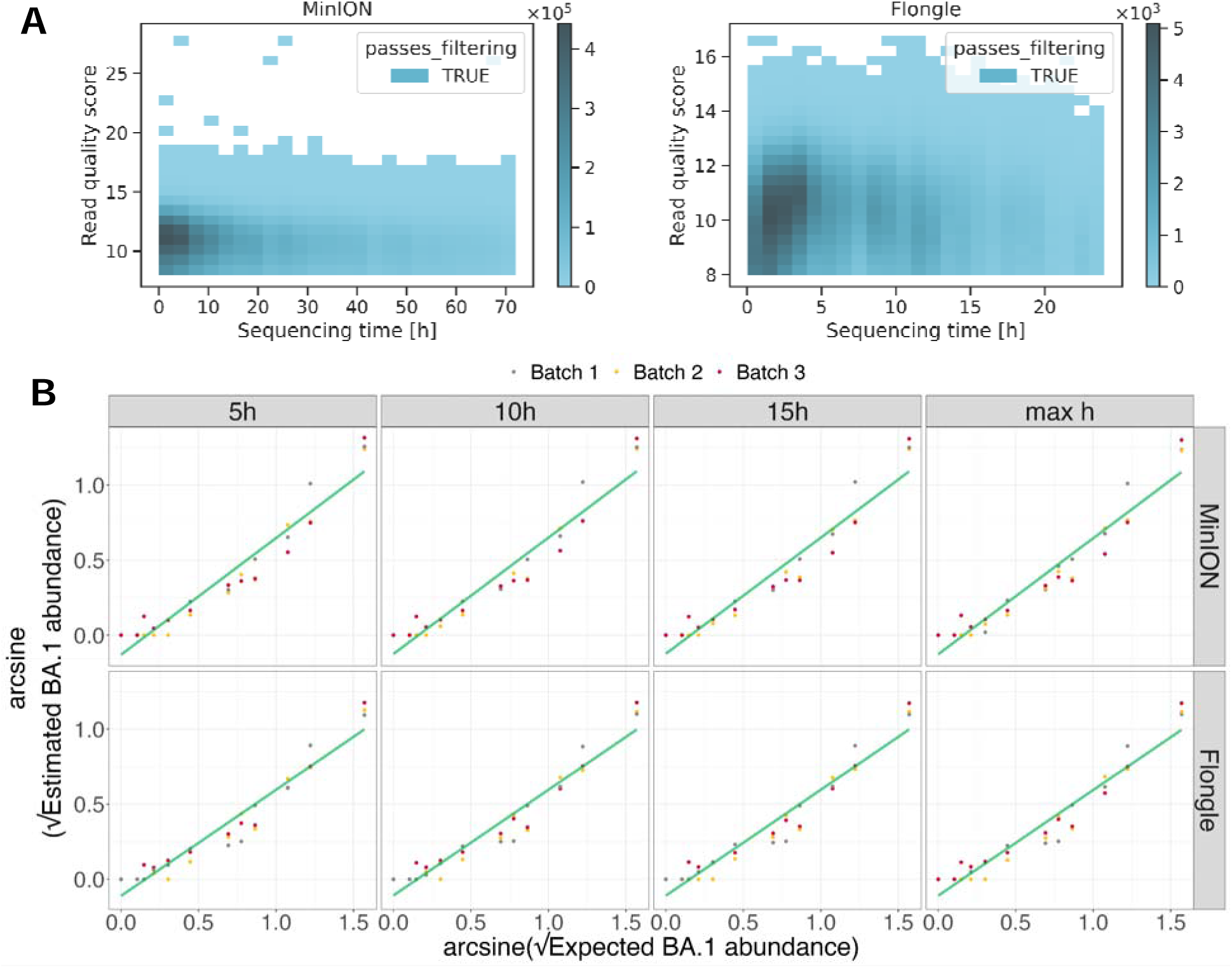
Variant abundance prediction for different runtimes of MinION and Flongle flow cell. **A:** Read sequence quality distribution by sequencing time. The read quality score is the mean base quality (Phred quality score) of a given read. Reads with a quality score below 8 are filtered out by the basecaller. **B:** Comparison of expected and estimated BA.1 abundance on arcsine-square root scale. The green line represents the derived linear model (see section 2.7).

## 4. Discussion

In this study, we investigated the suitability of different NGS technologies for sequencing and quantifying SARS-CoV-2 variants in wastewater. Our study adds to previous work comparing NGS technologies for clinical SARS-CoV-2 whole-genome analysis, in which both Illumina and MinION flow cell sequencing methods were effective, but varied in their error rates, quality of whole genome sequences, and detectability of short indels.^33^ Because wastewater is a unique matrix, distinct from clinical sampling due to the presence of inhibitory substances and co-occurrence of multiple circulating variants in a single sample, we benchmarked Illumina Novaseq 6000, Element Aviti, MinION flow cell, and Flongle flow cell sequencing techniques w.r.t. their ability to support wastewater-based epidemiology. Our findings indicate that, despite higher sequencing error rates and lower whole-genome coverage as compared to Illumina and Aviti, both MinION and Flongle flow cell sequencing demonstrate robust applicability to wastewater-based surveillance, due to variant abundance estimation being tolerant to sequencing errors. Thus, our findings suggest potential for improving efficiency, reducing cost, and increasing timeliness in genomic surveillance analyses.

The data generated with the novel Aviti sequencer showed the greatest concordance to Illumina sequencing data in terms of mutation frequencies and variant relative abundance estimates. Although the estimated variant abundances in some Aviti surveillance samples were misaligned, we showed that those were mainly associated with coverage drops under the v4.1 ARTIC primers used in this study^34^. The strong correlation of Aviti and Illumina-based measures is likely attributable to the low sequencing error rates of both systems. We found the Aviti sequencing error rate to be slightly lower than the Illumina sequencing error rate, which is consistent with first assessments of this novel technology^9^. Based on these findings, we conclude that Aviti sequencing poses a suitable alternative to Illumina sequencing in wastewater studies, especially in cases where sequencing quality is essential, such as detection of lowly abundant variants or variants characterised by a limited number of signature mutations.

Nanopore sequencing, and especially the R9.4.1 MinION flow cell, provides data of sufficient quality to allow classification of SARS-CoV-2 variants, albeit with weaker concordance to Illumina sequencing data than Aviti. In general, MinION and Illumina mutation frequencies showed good correlation, despite the higher sequencing error rate (0.1%) that we determined for the MinION flow cell. The MinION error rate determined in our study is slightly lower compared to the recently reported per-position error rate of 0.202% for R9.4.1 MinION flow cell sequencing on Twist synthetic RNA sequencing^35^. The lower rate observed here is likely due to the different source of SARS-CoV-2 RNA, as we used RNA from clinical samples as opposed to synthetic RNA. The higher error rate of nanopore sequencing impacted relative abundance estimates of variants, particularly for variants containing only a few signature mutations (e.g., BA.5 with only two signature mutations) as they are more susceptible to sequencing errors. Despite this challenge, the abundance estimates of the MinION surveillance samples correlated well with the Illumina-based estimates. We observed larger discrepancies of mutation frequency and variant abundance estimates between Flongle and Illumina surveillance data, which we attribute to the higher sequencing error rate of the Flongle flow cell, also observed by Mosbruger et al.^14^. The observed difference in sequencing error rate between the MinION and Flongle flow cell was surprising as they have the same underlying chemistry. However, Vereecke et al. previously reported differences in R9.4.1 MinION and R9.4.1 Flongle read quality and suggested that the discrepancy is due to deviations in *squiggle spaces*, the input of the base calling alogrithm^36^. In our study, the higher sequencing error rates of the nanopore flow cells resulted in an increased abundance of low-level spurious variants (noise) due to false or undefined nucleotide changes at genomic positions of variant-defining mutations. Analysis of the spike-in data revealed that the higher noise in the Nanopore data causes a significant underestimation of BA.1 abundance estimates compared to estimates from Illumina and Aviti sequencers. However, no significant difference in BA.1 abundance estimates was detected between the Nanopore flow cells, despite the previously reported higher error rate of Flongle flow cell sequencing. This is due to the larger number of signature mutations in BA.1^37^, making variant abundance estimation more robust to sequencing errors.

An advantage of nanopore sequencing-based technologies is the speed at which data can be generated, allowing for faster processing than both Illumina and Aviti sequencing^38^. Indeed, we showed by downsampling reads that reducing the runtime of MinION and Flongle flow cell sequencing to as little as five hours did not significantly change the derived abundance estimates, as for both flow cells a large proportion of good quality reads is produced within the first five hours of a sequencing run. These results imply substantial potential for reducing sequencing costs since reduced runtime enables the re-use of flow cells after subjecting them to a washing protocol as shown by Lipworth et al.^17^. Overall, our study suggests that the use of ONT flow cells is appropriate in scenarios where a prompt assessment of circulating variants has priority over achieving maximum precision in abundance estimation. This might be particularly beneficial in settings with higher cost and infrastructure restrictions where portable and cost-efficient sequencing technologies are prioritised.

The efficacy of sequencing technologies for wastewater is likely influenced by the number of signature mutations of the distinct circulating variants as well as the surveillance deconvolution timeframes. Both factors should therefore be considered when choosing an NGS method. In our study we had access to one week of wastewater surveillance data, but we believe that longer sampling time frames could positively impact variant abundance estimation. Since the variant deconvolution approach used in this study is tailored to time series data by allowing for kernel smoothing of variant abundance estimates over time^31^, we expect that smoothing over longer surveillance time periods would further decrease the divergence in variant abundance estimates between Illumina and Nanopore data. This prediction can be tested in the future by analysing data from longer time periods than one week. We also expect this effect to become more prominent as the number of signature mutations in the variant increases.

Another limitation of our study is that we did not assess the impact of longer sequencing reads, which, in principle, are available through Nanopore sequencing. It is expected that long-read whole-genome sequencing would increase the detection of co-occurring mutations on a single amplicon and hence improve variant abundance estimation. However, the extraction of long RNA fragments from wastewater covering the entire SARS-CoV-2 genome remains challenging^6^.

While our study emphasises NGS for WBE, it is also important to acknowledge that sequencing is just one element in the wastewater surveillance workflow. The limiting factors of implementing such workflow extend beyond the choice of sequencing technology, encompassing challenges in portable library preparation^39^, thermocycler accessibility^40^, and the need for sufficient computational resources for data analysis and abundance estimates. Here we used the V-pipe workflow to process the samples and subsequently calculate the SARS-CoV-2 abundance estimates ^30^. Although convenient to use, the workflow can require substantial computational resources and the use of a high performance computing cluster, especially for high-coverage samples. Efforts for reducing required resources and enabling real-time analysis will be crucial for advancing the accessibility of WBE methodologies.

## 5. Conclusion

Our study demonstrates that a diverse range of NGS technologies can be readily and reliably applied for wastewater surveillance studies. In terms of sequencing quality, the Element Aviti sequencer provides better sequencing data compared to Illumina. We also illustrated that Nanopore sequencing, especially the ONT MinION flow cell, resulted in similar mutation frequencies and variant abundance estimates as Illumina- and Aviti-based data despite lower sequencing quality. We further presented evidence that through nanopore real-time sequencing, the sequencing runtime can be reduced to five hours without changing variant abundance estimates significantly. MinION flow cell sequencing can be a valid sequencing method for wastewater studies where variants of interest are adequately abundant and have a sufficient number of signature mutations. For studies where the variants of interest can only be distinguished by a few mutations, Element Aviti sequencing can be a suitable alternative to Illumina sequencing. Together, these findings highlight that the selection of sequencing technology should be based on multiple factors, including circulating variants and associated mutations, cost, accessibility, and importance of timely mutation and variant abundance estimates.

## Supporting information

Supplementary Figures for Manuscript.pdf

## Data Availability

All data produced in the present work are made available and cited in the manuscript.

https://doi.org/10.3929/ethz-b-000663827

https://www.ebi.ac.uk/ena/browser/view/PRJEB44932

https://doi.org/10.5281/zenodo.11085721

## 6. Acknowledgements

Part of this work has been supported by the Swiss National Science Foundation [grant number No. CRSII5_205933] and by the Swiss Federal Office of Public Health. Further we would like to acknowledge Hai Bui, Adriana Hotz, Laura Neff, Kelsey Schaerer and Joel Wirz for the technical sequencing efforts done in the lab.

## 7. Declaration of generative AI and AI-assisted technologies in the writing process

During the preparation of this work the authors used ChatGPT in order to improve the readability of the text. After using this tool/service, the authors reviewed and edited the content as needed and take full responsibility for the content of the publication.

## 8. Ethics Statement

For the clinical isolates used in this study, ethical approval was waived by the Ethics Committee “Ethikkommission Nordwest- und Zentralschweiz (EKNZ)” in accordance with the Swiss Human Research Act (HRA) Art. 51(Project-ID Req-2020-00563).

