## Supplementary Figures for Manuscript.pdf for "Assessing different next-generation sequencing technologies for wastewater-based epidemiology"

(a)

(b)

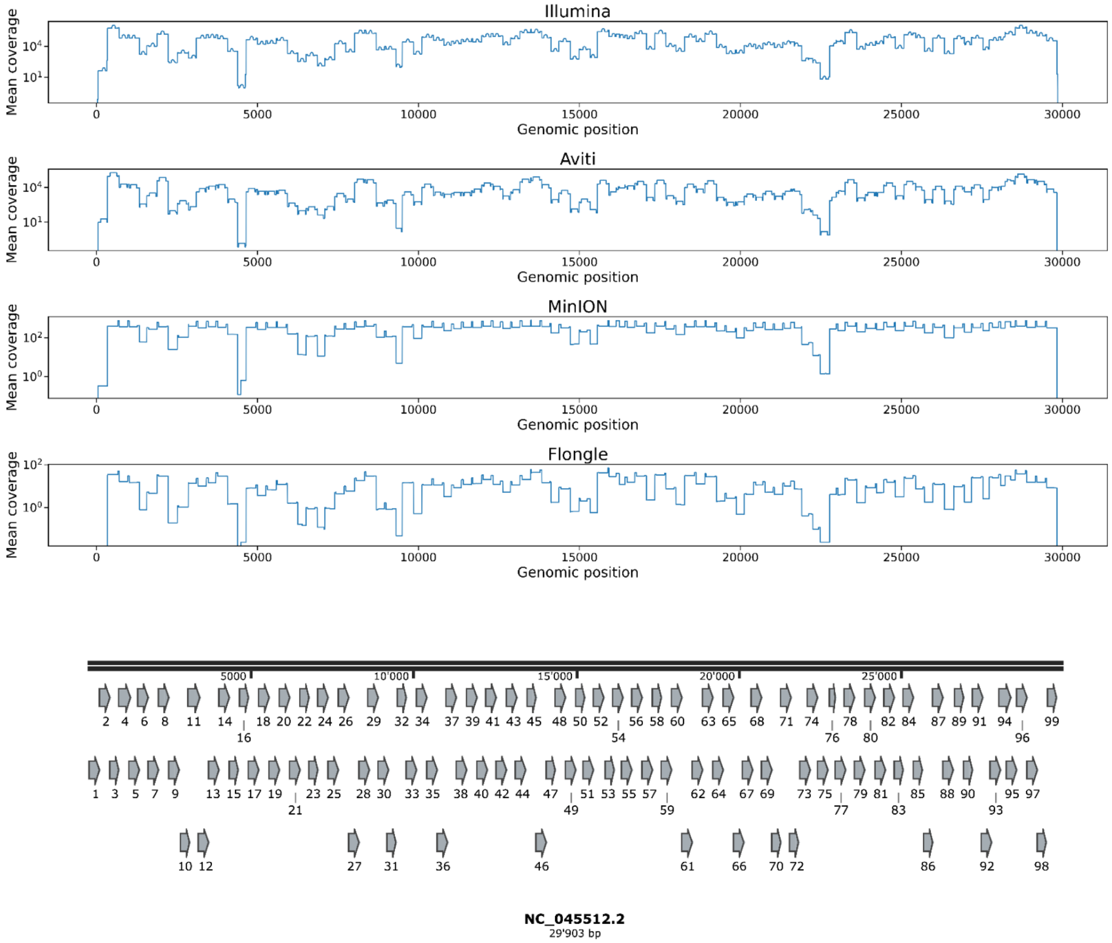


**Supplementary Figure A: (a)** Positional coverage of the SARS-Cov-2 (NC_045512.2) by sequencing technology. **(b)** Schematic representation of the distribution of the amplicons resulting from the v4.1 ARTIC primers over the SARS-Cov-2 genome (NC_045512.2).

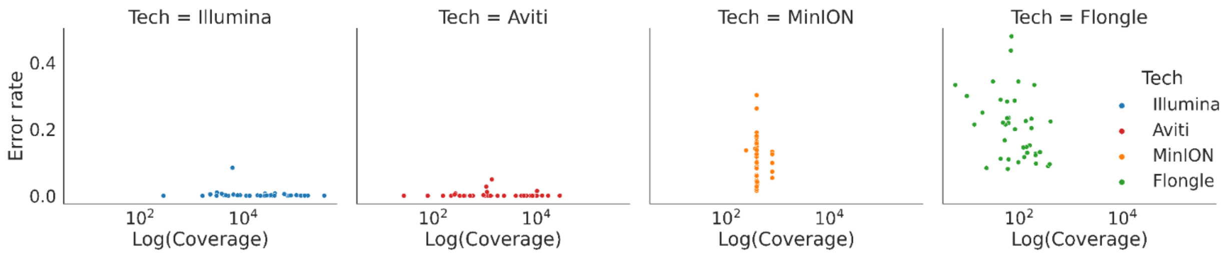


**Supplementary Figure B: Per position error rate by Log(Coverage).** Error rates were estimated from the non-diluted samples of the spike-in experiments, where the expected relative abundance of BA.1 is 1.0. Only positions corresponding to BA.1 signature mutations were considered. For more details see Methods.

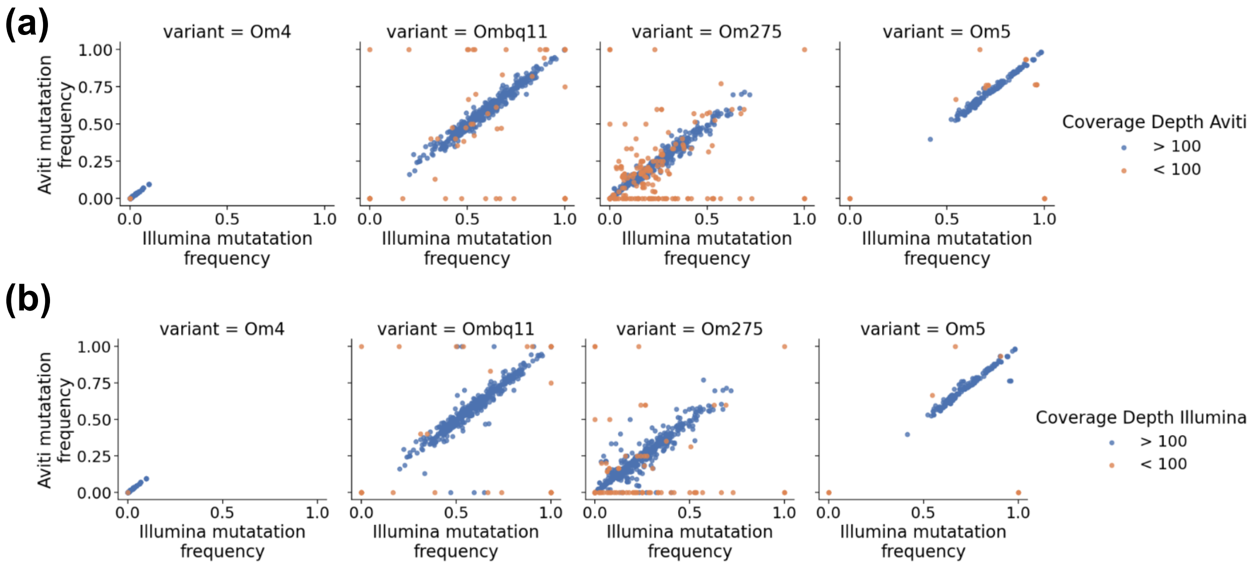


**Supplementary Figure C: Mutation frequencies of variant defining mutations compared between Illumina and Aviti by variant and coverage depth.** Each dot represents a unique mutation for the respective variant for one wastewater sample. Dots are coloured by (a) Aviti coverage depth or (b) Illumina coverage depth at the respective mutation position.

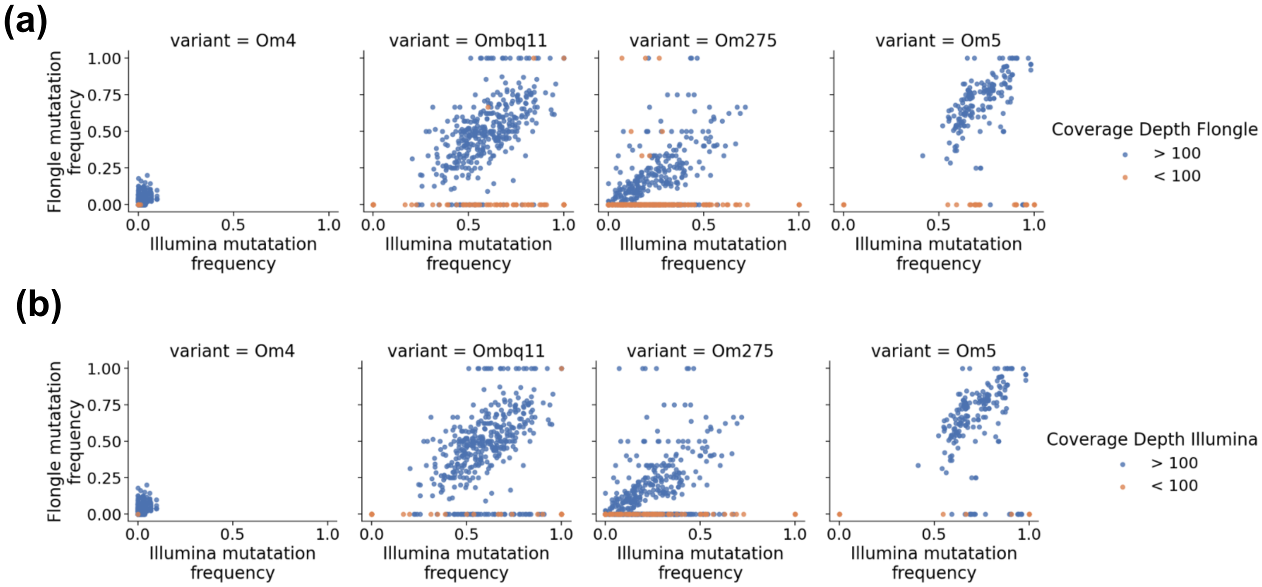


**Supplementary Figure D: Mutation frequencies of variant defining mutations compared between Illumina and MinION by variant and coverage depth.** Each dot represents a unique mutation for the respective variant for one wastewater sample. Dots are coloured by (a) MinION coverage depth or (b) Illumina coverage depth at the respective mutation position.

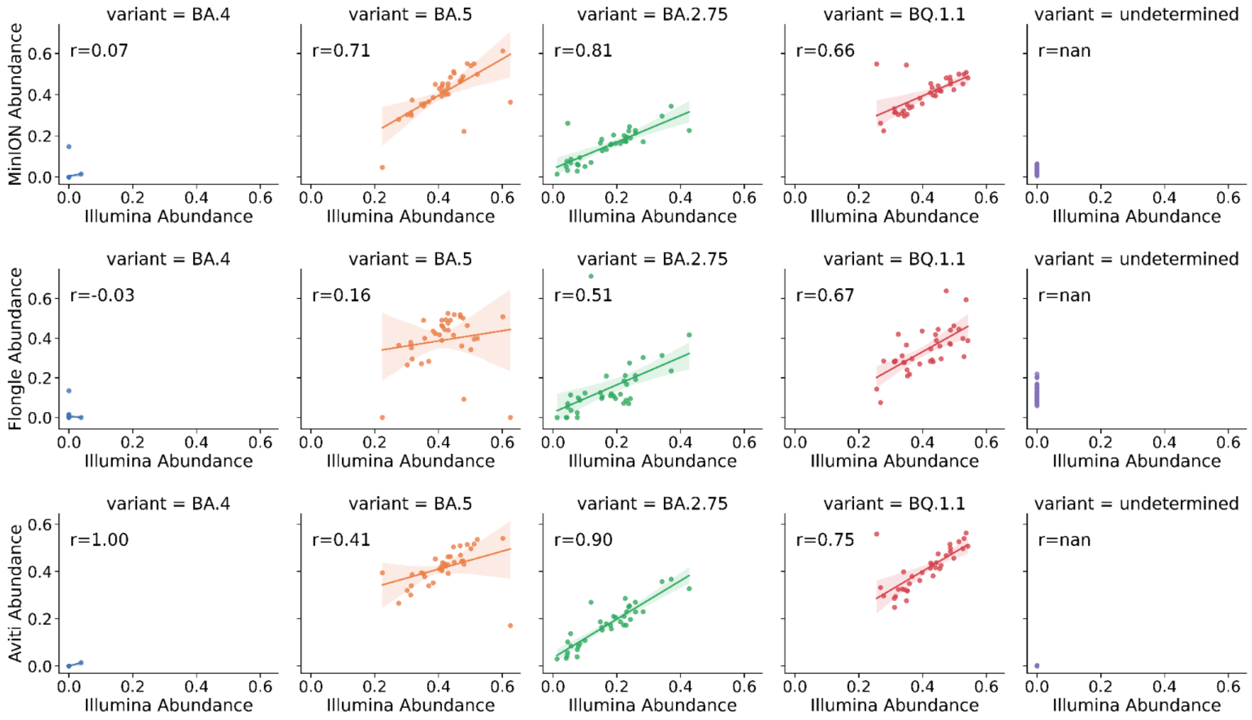


**Supplementary Figure E: Correlation of variant abundance estimations of the sequencing technologies of interest to Illumina-based variant abundance estimations by variant.** Each dot represents the estimated abundance of a given variant for one wastewater sample. The abundances were estimated using LolliPop. The r-squared values represent the $R^2$-statistic, deduced by squaring the respective Pearson correlation coefficient. The line through the points represents the estimated linear regression on the points and the shaded area the corresponding 95% confidence interval. Mutations that cannot be classified to a defined variant, are categorized as “undetermined” by LolliPop.

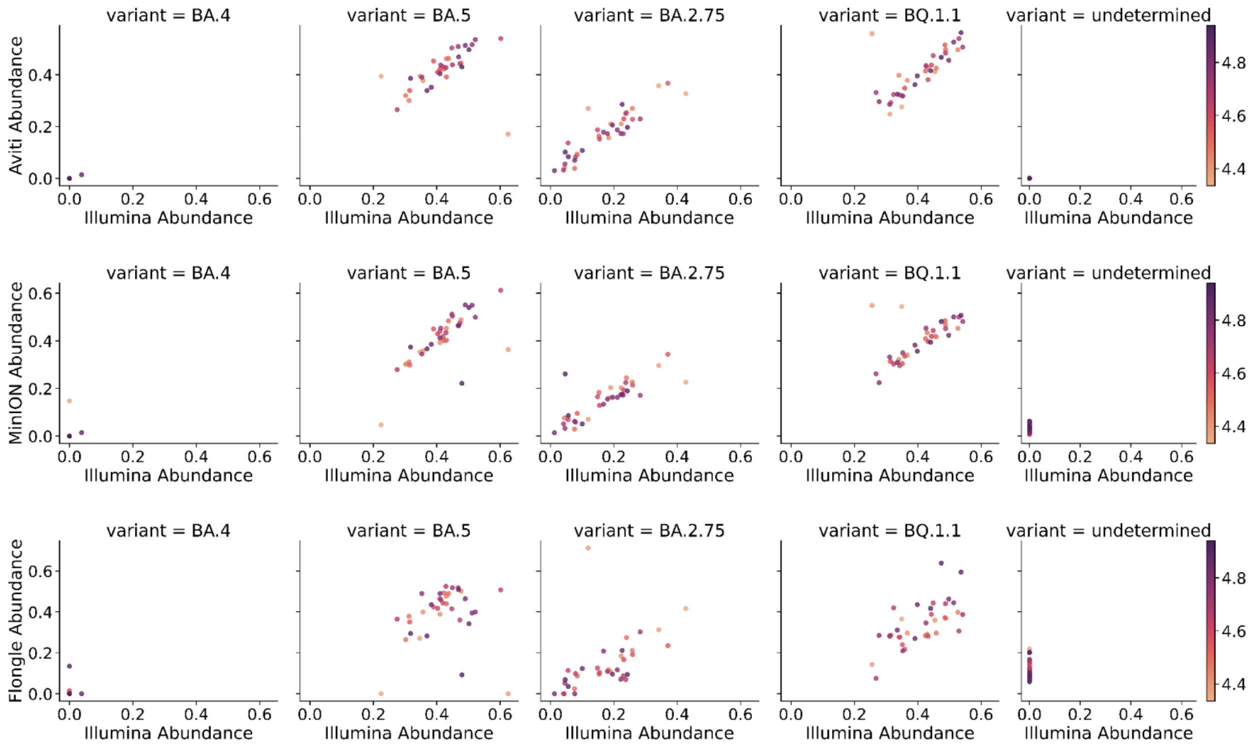


**Supplementary Figure F: Correlation of variant abundance estimations of the sequencing technologies of interest to Illumina-based variant abundance estimations by variant and mean sample coverage.** Each dot represents the estimated abundance of a given variant for one wastewater sample. Dots are coloured by the log(mean coverage) of the respective sample. Dark purple corresponds to a high coverage, light yellow to a low coverage. The abundances were estimated using LolliPop. Mutations that cannot be classified to a defined variant, are categorized as “undetermined” by LolliPop.

**(a)**

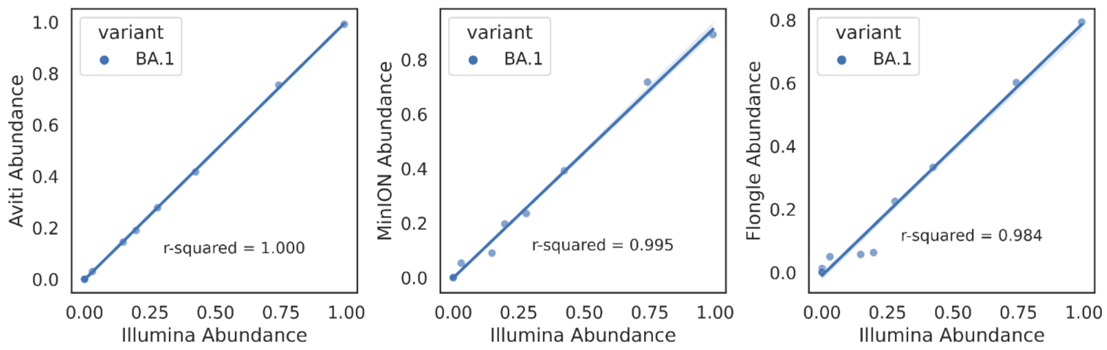


**(b)**

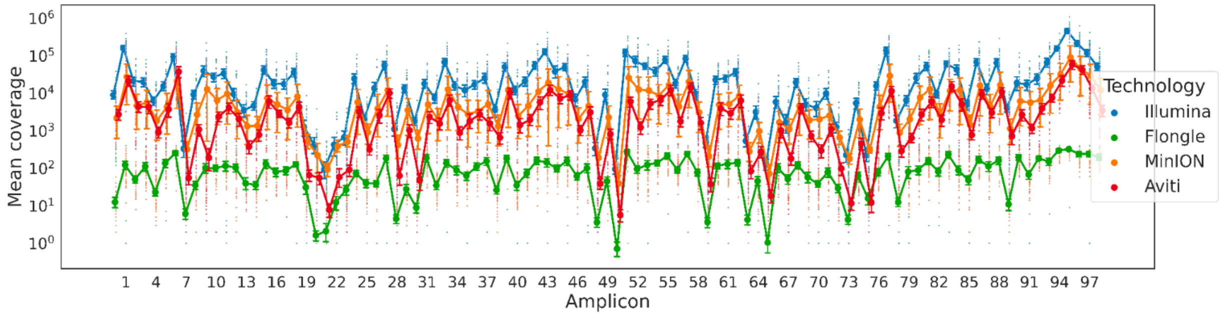


**Supplementary Figure G: (a) Comparison of variant abundance estimations by sequencing technologies of interest vs. Illumina.** Each dot represents the estimated abundance of a given variant for one wastewater sample. The abundances were estimated using LolliPop. The r-squared values represent the $R^2$-statistic, deduced by squaring the respective Pearson correlation coefficient. The line through the points represents the estimated linear regression on the points and the shaded area the corresponding 95% confidence interval. **(b) Mean amplicon coverage distribution for the sequencing technologies of interest.** The amplicons for all sequencing technologies were generated using v4.1 ARTIC primers. For each amplicon the mean coverage of the different sequencing technologies is represented by a large dot. The smaller dots represent the individual coverage depth of each nucleotide position of the amplicon. The vertical line at each amplicon represents the inter-quartile range of the coverage values (pi = 50).

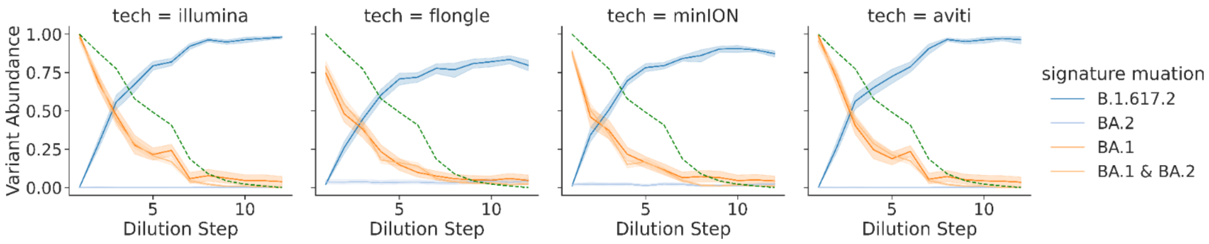


**Supplementary Figure H: Mutation frequency of BA.1 (Om1) defining mutations compared to expected BA.1 abundance.** For each sequencing technology, three technical replicates were provided. The bold line represents the mean abundance estimation of a given variant; the shaded area gives the 95% CI of the mean-values. The dotted green line shows the expected BA.1 concentration. Om2 = BA.2, delta = B.1.617.2.

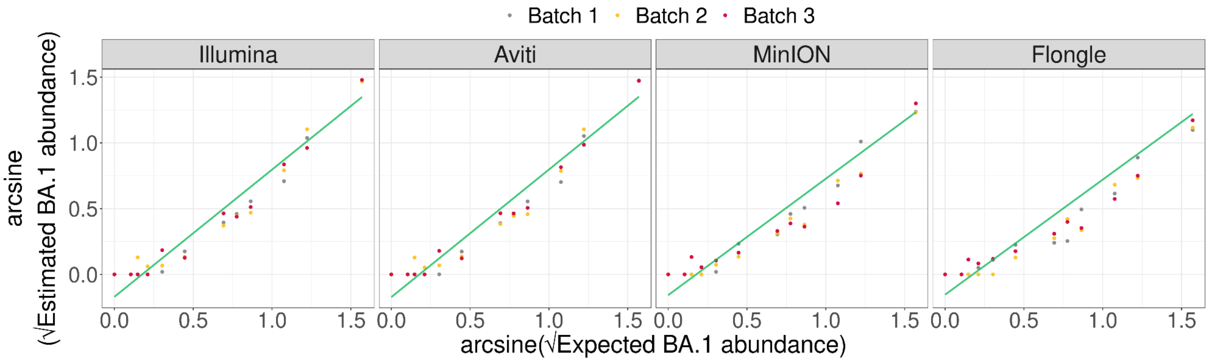


**Supplementary Figure I:** Comparison of expected and estimated BA.1 abundance for re-normalized data on arcsine-square root scale. The green line represents the derived linear model, for more information see Methods.

| | Coefficient | Estimate | Std. Error | t-value | p-value |
|---|---|---|---|---|---|
| **MinION (reference)** | intercept | -0.127254 | 0.026004 | -4.8936 | **2.758e-06** |
| | slope | 0.772821 | 0.045681 | 16.9177 | **< 2.2e-16** |
| **Illumina** | intercept | -0.031979 | 0.040441 | -0.7908 | 0.43046 |
| | slope | 0.153231 | 0.066049 | 2.3200 | **0.02183** |
| **Flongle** | intercept | 0.018164 | 0.034230 | 0.5306 | 0.59654 |
| | slope | -0.069811 | 0.058266 | -1.1981 | 0.23294 |
| **Aviti** | intercept | -0.033830 | 0.040613 | -0.8330 | 0.40632 |
| | slope | 0.154726 | 0.066051 | 2.3425 | **0.02060** |

**Supplementary Table A: Table stating linear model parameters with MinION as reference factor level determined from spike-in samples.** These parameters are used to display the response curves in Figure 5B. Significant levels were determined using a two-sided t-test.

| | Coefficient | Estimate | Std. Error | t-value | p-value |
|---|---|---|---|---|---|
| **Flongle (reference)** | intercept | -0.109091 | 0.022259 | -4.9009 | **2.672e-06** |
| | slope | 0.703010 | 0.036169 | 19.4370 | **< 2.2e-16** |
| **Illumina** | intercept | -0.050143 | 0.038141 | -1.3147 | 0.1908381 |
| | slope | 0.223043 | 0.059865 | 3.7257 | **0.0002846** |
| **MinION** | intercept | -0.018164 | 0.034230 | -0.5306 | 0.5965397 |
| | slope | 0.069811 | 0.058266 | 1.1981 | 0.2329437 |
| **Aviti** | intercept | -0.051993 | 0.038323 | -1.3567 | 0.1771243 |
| | slope | 0.224537 | 0.059868 | 3.7506 | **0.0002602** |

**Supplementary Table B: Table stating linear model parameters with Flongle as reference factor level determined from spike-in samples.** Significant levels were determined using a two-sided t-test.

| | Coefficient | Estimate | Std. Error | t-value | p-value |
|---|---|---|---|---|---|
| **MinION (reference)** | intercept | -0.1576217 | 0.0329275 | -4.7869 | **4.359e-06** |
| | slope | 0.8851541 | 0.0610747 | 14.4930 | **< 2.2e-16** |
| **Illumina** | intercept | -0.0128718 | 0.0473792 | -0.2717 | 0.7863 |
| | slope | 0.0821092 | 0.0826288 | 0.9937 | 0.3221 |
| **Flongle** | intercept | 0.0035631 | 0.0465183 | 0.0766 | 0.9391 |
| | slope | -0.0103122 | 0.0854885 | -0.1206 | 0.9042 |
| **Aviti** | intercept | -0.015302 | 0.0475985 | -0.3215 | 0.7483 |
| | slope | 0.0850477 | 0.0827049 | 1.0283 | 0.3056 |

**Supplementary Table C: Table stating linear model parameters with MinION as reference factor level determined from re-normalised spike-in samples.** These parameters are used to display the response curves in Supplementary Figure E. Significant levels were determined using a two-sided t-test.

|  | Coefficient | Estimate | Std. Error | t-value | p-value |
|---|---|---|---|---|---|
| **Flongle (reference)** | intercept | -0.1540585 | 0.0328593 | -4.6884 | **6.613e-06** |
|  | slope | 0.8748419 | 0.0598178 | 14.6251 | **< 2.2e-16** |
| **Illumina** | intercept | -0.0164350 | 0.0473319 | -0.3472 | 0.7290 |
|  | slope | 0.0924214 | 0.0817042 | 1.1312 | 0.2600 |
| **MinION** | intercept | -0.0035631 | 0.0465183 | -0.0766 | 0.9391 |
|  | slope | 0.0103122 | 0.0854885 | 0.1206 | 0.9042 |
| **Aviti** | intercept | -0.0188652 | 0.0475514 | -0.3967 | 0.6922 |
|  | slope | 0.0953599 | 0.0817811 | 1.1660 | 0.2456 |

**Supplementary Table D: Table stating linear model parameters with Flongle as reference factor level determined from re-normalised spike-in samples.** Significant levels were determined using a two-sided t-test.

| MinION | Coefficient | Estimate | Std. Error | t-value | p-value | Number of ‘passed’ reads |
|---|---|---|---|---|---|---|
| **5 h (reference)** | intercept | -0.130156 | 0.031772 | -4.2586 | **4.027e-05** | 3747044 |
| | slope | 0.7770553 | 0.0500585 | 15.5230 | **<2.2e-16** | |
| **10 h** | intercept | 0.0050643 | 0.0421691 | 0.1201 | 0.9046 | 2631387 |
| | slope | -0.0023326 | 0.0698824 | -0.0334 | 0.9734 | |
| **15 h** | intercept | 0.0066611 | 0.0418917 | 0.1590 | 0.8739 | 1819991 |
| | slope | -0.0041478 | 0.0697517 | -0.0595 | 0.9527 | |
| **max h** | intercept | 0.0029020 | 0.0401504 | 0.0723 | 0.9425 | 7051031 |
| | slope | -0.0042343 | 0.0678083 | -0.0624 | 0.9503 | |

**Supplementary Table E: Table stating linear model parameters with 5 h run-time as reference factor level determined from MinION time subsampled spike-in samples.** Significant levels were determined using a two-sided t-test.

| Flongle | Coefficient | Estimate | Std. Error | t-value | p-value | Number of 'passed' reads |
|---|---|---|---|---|---|---|
| **5 h (reference)** | intercept | -0.1106250 | 0.0305631 | -3.6196 | **0.0004152** | 215592 |
| | slope | 0.7059255 | 0.0500585 | 14.1020 | **< 2.2e-1** | |
| **10 h** | intercept | 0.0028492 | 0.0421691 | 0.0676 | 0.9462311 | 99593 |
| | slope | -0.0026447 | 0.0698824 | -0.0378 | 0.9698670 | |
| **15 h** | intercept | 0.0015978 | 0.0418917 | 0.0381 | 0.9696303 | 69256 |
| | slope | -0.0012549 | 0.0697517 | -0.0180 | 0.9856723 | |
| **max h** | intercept | 0.0015343 | 0.0401504 | 0.0382 | 0.9695737 | 38752 |
| | slope | -0.0029159 | 0.0678083 | -0.0430 | 0.9657630 | |

**Supplementary Table F: Table stating linear model parameters with 5 h run-time as reference factor level determined from Flongle time subsampled spike-in samples.** Significant levels were determined using a two-sided t-test.
